# Adjusted booster schedules disperse age-dependent differences in antibody titers benefitting risk populations

**DOI:** 10.1101/2022.08.08.22278545

**Authors:** Lisa Müller, Marcel Andrée, Wiebke Moskorz, Ingo Drexler, Sandra Hauka, Johannes Ptok, Lara Walotka, Ramona Grothmann, Jonas Hillebrandt, Anastasia Ritchie, Laura Peter, Andreas Walker, Jörg Timm, Ortwin Adams, Heiner Schaal

## Abstract

We provide follow-up data on the humoral immune response after COVID-19 vaccinations of a cohort aged below 60 and over 80 years. While anti-SARS-CoV-2 spike-specific IgG and neutralization capacity waned rapidly after initial vaccination, additional boosters highly benefitted humoral immune responses including neutralization of Omikron variants in the elderly cohort.

## Introduction

From the onset of the SARS-CoV-2 pandemic in 2019, it quickly became clear that only prophylactic immunization offered a way out of this global health crisis [1, 2]. In the following year 2020, several manufacturers received emergency use authorization for their vaccines against SARS-CoV-2. This included the novel class of mRNA vaccines, Comirnaty (BioNTech/Pfizer) and Spikevax/mRNA-1273 (Moderna) [3, 4].

In Germany and other countries, immunization schedules were rolled out at the end of December 2020, prioritizing risk populations including immunocompromised and elderly individuals [5]. With mRNA vaccines being the first of their kind, vaccinated risk populations were in the focus of monitoring studies to evaluate the magnitude and quality of the immune response to these vaccines [6, 7]. While initially, both mRNA formulations were designed as “prime and boost” immunizations, several studies quickly pointed out the potential necessity of at least a third vaccination, especially for populations with a generally reduced or short lived humoral immune response [2, 8].

With the emergence of more variants of concern such the currently predominant Omikron variants which again caused a sharp surge in cases worldwide, the necessity of adjusted vaccination schedules became more evident. Therefore, less than a year after the first vaccination campaigns started, several countries began their additional booster campaigns. However, the combination of increasing case numbers and both, hetero- and homologous booster vaccinations, predominantly using mRNA vaccines, has led to a complex mixture of SARS-CoV-2-specific immunological profiles throughout the population.

Early in 2021, we performed a cohort study with two distinct age groups, vaccinees below 60 and above 80 years [9]. Here, we present a follow up of this vaccinee cohort. We revisited the study cohort half a year and one year after their initial prime and boost vaccination. We monitored the cohort for their total anti-SARS-CoV-2 spike-specific and nucleocapsid-specific immune response as well as the magnitude of the neutralizing antibody response. Now that Omikron is the most prevalent variant, we also included the neutralizing antibody response to the Omikron BA.1 and BA.5. The follow-up of this cohort provides an important snapshot of current immunological profiles.

## Results

Participants were volunteers from the SBK nursing home in Cologne, Germany who participated in both blood samplings for first part of the study (blood collection BC#1 and BC#2, analyzed in Müller et. al., 2021 [9]) and in both follow-ups (blood collection BC#3 and BC#4) and gave informed consent (n = 84). They received their 1^st^ vaccination with the BioNTech/Pfizer vaccine at the end of December 2020 and their 2^nd^ in late January 2021. All individuals received their 3^rd^ vaccination (97.7% BioNTech, 2.3% Moderna) between September and November 2021. Additionally, 39 (46.4 %) of the participants received a 4^th^ vaccination in early February 2022 (100% BioNTech), 8 of the younger vaccinees, 31 of the elderly.

Blood samples were analyzed for anti-SARS-CoV-2 nucleocapsid-specific (Abbott) and spike-specific (Euroimmun) IgG titers and neutralizing SARS-CoV-2 antibodies against B.1 WT (EPI_ISL_425126), Omikron BA.1 (EPI_ISL_12813299.1) and BA.5 (EPI_ISL_14167576) isolates about 6- and 12-months (BC#3 and BC#4) after their first vaccination to screen for age-related differences in the longevity of the humoral immune response and the induction of the antibody response post booster-vaccinations. Participants who reported an infection six months prior to the second follow-up (n =20) were analyzed separately. This resulted in a final cohort of 64 vaccinated individuals (28 younger, 36 elderly).

At the first follow-up blood collection (BC#3) six months after their first vaccination, the quantitative SARS-CoV-2 spike-specific IgG titers again differed significantly between the two groups (p = 0.0435). For the group of elderly vaccinees, the mean IgG titer was 99.98 BAU/mL, ranging from 3.45 to 1111.0 BAU/mL. In this group, 41.6% of the tested individuals had titers below cut-off (>35.2 BAU/mL). In the younger cohort, IgG titers ranged from 84.3 to 823.5 BAU/ml with a mean of 231.6 BAU/ml with no participants testing below cut-off (Figure 1A). At the second follow-up (BC#4) more than one year after the initial vaccination schedules, all participants had received a 3^rd^ vaccination. Furthermore, 86% (n= 31) of the elderly participants received a 4^th^ vaccination, while only 28% (n = 8) of the younger participants received the additional booster. At this blood collection (BC#4), SARS-CoV-2 spike-specific IgG titers were comparable between the age groups. Interestingly, the mean SARS-CoV-2 spike-specific IgG titer in the group of elderly vaccinees was two-fold higher (3337 BAU/mL) than for younger vaccinees (1663 BAU/mL) and no participant tested below cut-off. However, titers ranged wider in the former group (68.0 – 10800 BAU/mL) than in the latter (300 – 6950 BAU/mL).

**Figure 1:**
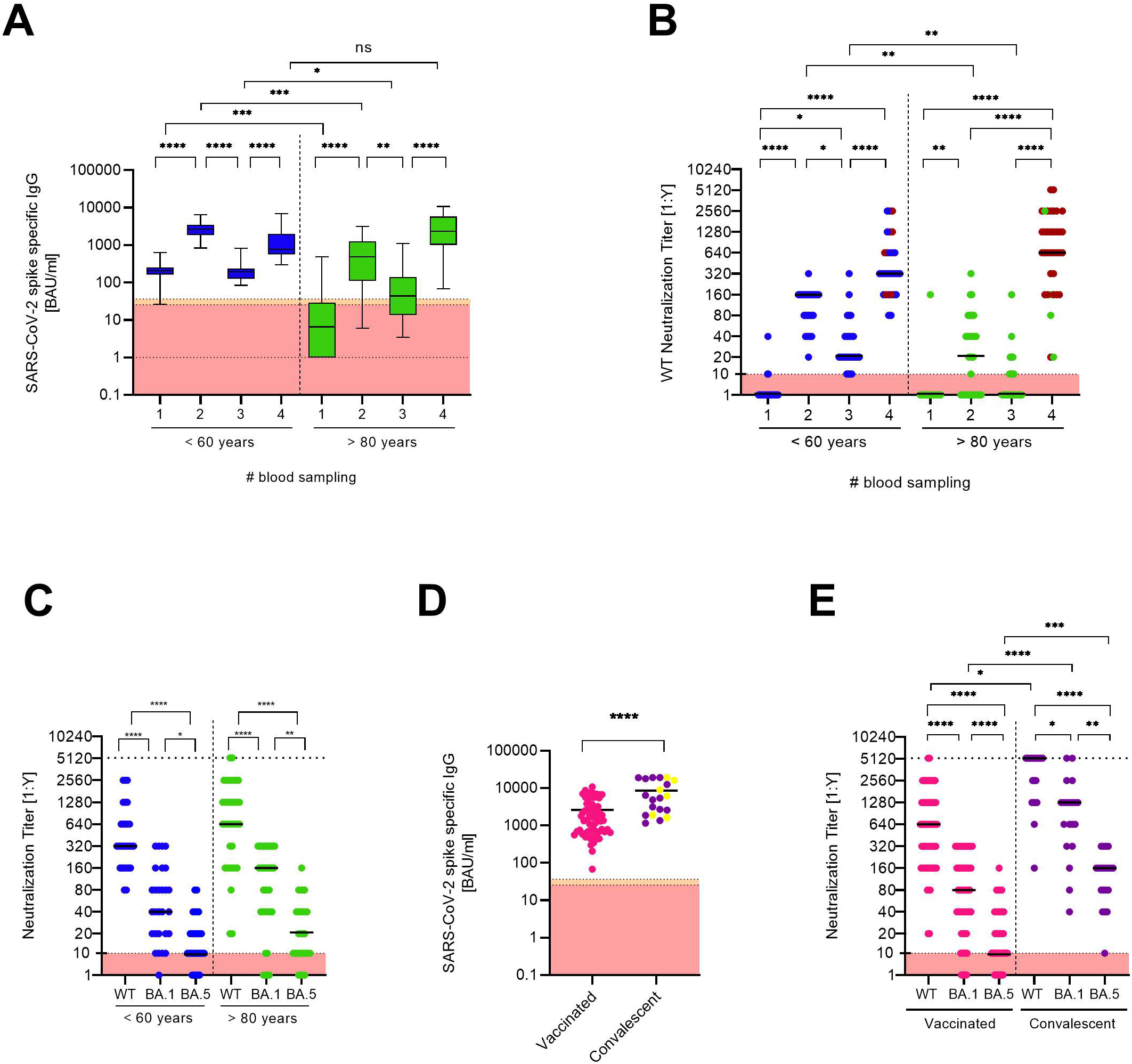
**A** Anti-SARS-CoV-2 spike specific antibody titers were determined for both age groups at each blood collection (BC#1 and BC#2 analyzed in Müller et. al., 2021 [9]). Antibody titers below the detection limit were set to 1.0. Boxes span the interquartile range; the line within each box denotes the median and whiskers indicate the 2.5 and 97.5 percentile values. **B** Neutralization titers of the two age groups were measured at each blood collection. Individuals who received an additional booster vaccination before the fourth blood collection are indicated with red symbols. **C** Comparison of the neutralization titers against a B.1 WT strain and Omikron BA.1 and BA.5 isolates between the two age groups at the fourth blood collection. **D** Comparison of anti-SARS-CoV-2 spike IgG titers between vaccinated only and vaccinated convalescent individuals at the fourth blood collection. Convalescent individuals with an anti-SARS-CoV-2 nucleocapsid-specific IgG titer considered positive are marked in yellow. **E** Comparison of the neutralization titers against B.1 WT and BA.1 and BA.5 Omikron isolates of the vaccinated only and vaccinated convalescent cohort. All data sets were tested with the Shapiro-Wilk test for normality. As most data sets did not show normal Gaussian distribution, parametric tests were performed. Two unpaired data sets (D) were compared by two-tailed Mann Whitney test. Comparison of two paired data sets (C and E) were done by two-tailed Wilcoxon matched-pairs signed rank test. For the comparison of antibody titers at different time points (A and B), tests within age groups were tested with two-tailed Friedman Test and comparison between age groups were tested with Kruskal Wallis Test (between), both followed by Dunn’ s multiple comparison test. All tests were performed using GraphPad Prism software Version 9.3.1.

Furthermore, we compared the time-dependent progress of the neutralization capacity against a SARS-CoV-2 B.1 WT isolate between the age groups (Figure 1B). At the first follow-up (BC#3), the median neutralization titer (MNT) in the group of elderly participants drastically decreased to 0. After the elderly participants received their 3^rd^ and 4^th^ vaccination (BC#4), however, the MNT for the group of elderly participants was 640. For the group of younger vaccinees, MNT were significantly higher compared to the elderly. At the first follow-up (BC#3), median neutralization capacity against the B.1 WT isolate in this group was decreased to 20. At the one-year follow-up blood collection (BC#4), the MNT in the group of younger vaccinees was 320.

The neutralization capacity determined by MNT was significantly decreased against both Omikron variants (BA.1 and BA.5) in both groups as compared to the B.1 WT. The MNT against Omikron BA.1 in the group of elderly participants was 160, against BA.5 it was decreased to 20 compared to 640 against the WT. Younger vaccinees showed an MNT of 40 against BA.1 and 10 against BA.5 compared to 320 against the WT isolate (Figure 1C).

Furthermore, we separately analyzed the group of vaccinated convalescent participants (n = 20) compared to the overall vaccinated cohort (n = 64). The 20 convalescent participants (10 younger and 10 older vaccinees), reported an infection confirmed by PCR within the past six months prior to the second follow-up blood collection, 9 were infected in a community outbreak in early February 2022. A comparison of overall anti-SARS-CoV-2 spike-specific IgG titers showed significantly higher titers (p <0.0001) in the group of convalescent participants with a mean titer of 8560 BAU/mL compared to 2605 BAU/mL in the vaccinated cohort. It is of note that only 6 of 20 convalescent participants had anti-SARS-CoV-2 nucleocapsid-specific titers considered positive (Figure 1D).

In this cohort, we also analyzed neutralizing antibodies and compared the WT and Omikron titers against the vaccinated cohort. In line with overall anti-SARS-CoV-2 spike-specific IgG levels, the convalescent cohort displayed significantly higher neutralizing antibody levels against WT and both Omikron isolates. However, despite the majority being infected with Omikron during the community outbreak in February 2022, Omikron neutralization titers were still lower than WT titers (Figure 1E).

## Discussion

We present the follow-up analysis of the SARS-CoV-2-specific antibody response to the BioNTech/Pfizer prime/boost vaccination of a cohort consisting of two age groups half a year and one year after their first COVID-19-vaccination.

Both mRNA vaccinations that received emergency approval in late 2020 were initially designed as “prime/boost” vaccination schedules, however, longitudinal effects of the vaccinations were still to be determined. While it is evident that prophylactic immunizations decreased the pandemic burden and positively influenced the development of hospitalization and death rates [10], various studies pointed out potential limitations of the COVID-19-vaccinations in specific sub-cohorts. These include immunocompromised patients, in particular organ transplant recipients [11] as well as elderly where the reduced induction of the humoral immune response after vaccination can likely be attributed to the effect of immunosenescence.

In this follow-up study, we observed a drastic decrease of the anti-SARS-CoV-2 spike-specific IgG titer at the first follow-up blood collection six months after the cohort received their first vaccination. Especially in the group of elderly participants, these effects were even more pronounced, with more than a third of the elderly vaccinees testing below cut-off. This is in line with other studies which report a rapid antibody waning after prime/boost regimens [12]. It further underlines the necessity of strategies to overcome such limitations with additional boosters.

Thus, shortly after the first follow-up, study participants received an asynchronous 3^rd^ mRNA vaccination between September and November 2021, less than a year after their first vaccination. In early February 2022, Germany’s vaccination commission recommended another booster for risk groups, so the majority of elderly participants received a 4^th^ vaccination before the second follow-up blood collection. This fully dispersed the age-dependent difference in mean IgG titers and neutralization capacity, although, as previously described, neutralization capacity against the Omikron BA.1 was drastically reduced, which was even more pronounced for BA.5 [13]. As shown in large cohort studies with participants aged over 60, the 4^th^ vaccination resulted in elevated protection from severe illness, hospitalization and death and even short-lived protection from infection and thus, proved highly beneficial for risk groups [14, 15].

In the group of convalescent individuals, the humoral immune response was significantly increased compared to the vaccinated cohort, which is in line with previous results [16]. While a high number of convalescents was infected during an Omikron BA.1 outbreak shortly before the second follow-up blood collection, both Omikron neutralization titers were lower than the neutralization titers against the B.1 WT, although neutralization titers against the WT were also significantly increased, suggesting a simultaneous induction of cross-reactive neutralizing antibodies against both variants [17]. Furthermore, anti-SARS-CoV-2 nucleocapsid-specific antibodies traditionally used as serological marker for natural infections were only detected in the minority of convalescent individuals. With increasing break-through infections in vaccinated individuals and data from large-scale cohort studies that showed the rapid decline of nucleocapsid specific antibodies especially in vaccinees [18], it seems highly likely that this serological marker will become even more unreliable with the rise of more complex immunological profiles.

## Data Availability

All data produced in the present study are available upon reasonable request to the authors.

## Notes

### Ethics Approval

The ethics committee of the Medical Faculty at the Heinrich-Heine University Düsseldorf, Germany (study no. 2021-1287), approved the study.

## Acknowledgments

The authors thank the members of Sozial Betriebe Köln, SBK for excellent organization and support and for the opportunity to conduct this study and all helpers and the caregivers at SBK for their support at the blood collection days. We would like to thank all volunteers at the SBK nursing home. We thank all members of the Virology department and especially Yvonne Dickschen, Alexandra Graupner and Björn Wefers for their excellent technical assistance.

## Author contributions

Conceptualization: HS, OA, MA, LM, Formal analysis: WM, LM, Investigation: All Authors; Writing – original draft preparation: HS, WM, OA, MA, LM, Writing – review and editing: All Authors, Supervision: HS, OA, MA, LM

## References

1. Viana J, van Dorp CH, Nunes A, et al. Controlling the pandemic during the SARS-CoV-2 vaccination rollout. Nat Commun 2021; 12(1): 3674.

2. Dagan N, Barda N, Kepten E, et al. BNT162b2 mRNA Covid-19 Vaccine in a Nationwide Mass Vaccination Setting. N Engl J Med 2021; 384(15): 1412–23.

3. Baden LR, El Sahly HM, Essink B, et al. Efficacy and Safety of the mRNA-1273 SARS-CoV-2 Vaccine. N Engl J Med 2021; 384(5): 403–16.

4. Polack FP, Thomas SJ, Kitchin N, et al. Safety and Efficacy of the BNT162b2 mRNA Covid-19 Vaccine. N Engl J Med 2020; 383(27): 2603–15.

5. Cylus J, Panteli D, van Ginneken E. Who should be vaccinated first? Comparing vaccine prioritization strategies in Israel and European countries using the Covid-19 Health System Response Monitor. Isr J Health Policy Res 2021; 10(1): 16.

6. Oliveira-Silva J, Reis T, Lopes C, et al. Humoral response to the SARS-CoV-2 BNT162b2 mRNA vaccine: Real-world data from a large cohort of healthcare workers. Vaccine 2022; 40(4): 650–5.

7. Swift MD, Breeher LE, Tande AJ, et al. Effectiveness of Messenger RNA Coronavirus Disease 2019 (COVID-19) Vaccines Against Severe Acute Respiratory Syndrome Coronavirus 2 (SARS-CoV-2) Infection in a Cohort of Healthcare Personnel. Clinical infectious diseases : an official publication of the Infectious Diseases Society of America 2021; 73(6): e1376–e9.

8. Wieske L, van Dam Kpj, Steenhuis M, et al. Humoral responses after second and third SARS-CoV-2 vaccination in patients with immune-mediated inflammatory disorders on immunosuppressants: a cohort study. Lancet Rheumatol 2022; 4(5): e338–e50.

9. Müller L, Andree M, Moskorz W, et al. Age-dependent Immune Response to the Biontech/Pfizer BNT162b2 Coronavirus Disease 2019 Vaccination. Clinical infectious diseases : an official publication of the Infectious Diseases Society of America 2021; 73(11): 2065–72.

10. Moghadas SM, Vilches TN, Zhang K, et al. The impact of vaccination on COVID-19 outbreaks in the United States. medRxiv 2021.

11. Lee A, Wong SY, Chai LYA, et al. Efficacy of covid-19 vaccines in immunocompromised patients: systematic review and meta-analysis. Bmj 2022; 376: e068632.

12. Shrotri M, Navaratnam AMD, Nguyen V, et al. Spike-antibody waning after second dose of BNT162b2 or ChAdOx1. Lancet 2021; 398(10298): 385–7.

13. Hachmann NP, Miller J, Collier AY, et al. Neutralization Escape by SARS-CoV-2 Omicron Subvariants BA.2.12.1, BA.4, and BA.5. N Engl J Med 2022; 387(1): 86–8.

14. Bar-On YM, Goldberg Y, Mandel M, et al. Protection by a Fourth Dose of BNT162b2 against Omicron in Israel. N Engl J Med 2022; 386(18): 1712–20.

15. Arbel R, Sergienko R, Friger M, et al. Effectiveness of a second BNT162b2 booster vaccine against hospitalization and death from COVID-19 in adults aged over 60 years. Nature medicine 2022; 28(7): 1486–90.

16. Muller L, Andree M, Ostermann PN, et al. SARS-CoV-2 Infection in Fully Vaccinated Individuals of Old Age Strongly Boosts the Humoral Immune Response. Front Med (Lausanne) 2021; 8: 746644.

17. Garcia-Beltran WF, St Denis KJ, Hoelzemer A, et al. mRNA-based COVID-19 vaccine boosters induce neutralizing immunity against SARS-CoV-2 Omicron variant. medRxiv 2021.

18. Krutikov M, Palmer T, Tut G, et al. Prevalence and duration of detectable SARS-CoV-2 nucleocapsid antibodies in staff and residents of long-term care facilities over the first year of the pandemic (VIVALDI study): prospective cohort study in England. Lancet Healthy Longev 2022; 3(1): e13–e21.

